# Racial/Ethnic, Biomedical, and Sociodemographic Risk Factors for COVID-19 Positivity and Hospitalization in the San Francisco Bay Area

**DOI:** 10.1101/2022.04.03.22273345

**Authors:** Wendy K. Tam Cho, David G. Hwang

## Abstract

**BACKGROUND:** The COVID-19 pandemic has uncovered clinically meaningful racial/ethnic disparities in COVID-19-related health outcomes. Current understanding of the basis for such an observation remains incomplete, with both biomedical and social/contextual variables proposed as potential factors.

**PURPOSE:** Using a logistic regression model, we examined the relative contributions of race/ethnicity, biomedical, and socioeconomic factors to COVID-19 test positivity and hospitalization rates in a large academic health care system in the San Francisco Bay Area prior to the advent of vaccination and other pharmaceutical interventions for COVID-19.

**RESULTS:** Whereas socioeconomic factors, particularly those contributing to increased social vulnerability, were associated with test positivity for COVID-19, biomedical factors and disease co-morbidities were the major factors associated with increased risk of COVID-19 hospitalization. Hispanic individuals had a higher rate of COVID-19 positivity, while Asian persons had higher rates of COVID-19 hospitalization. Diabetes was an important risk factor for COVID-19 hospitalization, particularly among Asian patients, for whom diabetes tended to be more frequently undiagnosed and higher in severity.

**CONCLUSIONS:** We observed that biomedical, racial/ethnic, and socioeconomic factors all contributed in varying but distinct ways to COVID-19 test positivity and hospitalization rates in a large, multiracial, socioeconomically diverse metropolitan area of the United States. The impact of a number of these factors differed according to race/ethnicity. Improving overall COVID-19 health outcomes and addressing racial and ethnic disparities in COVID-19 outcomes will likely require a comprehensive approach that incorporates strategies that target both individual-specific and group contextual factors.

## Introduction

As a discrete, novel, and large exogenous health shock, a pandemic can be viewed as a “natural experiment” that can yield manifold insights into disease transmission dynamics, population health status and equity, and health care system responsiveness, unfettered by pre-existing differences and structural inequalities associated with already prevalent diseases. The COVID-19 pandemic presents such a unique opportunity at a time when detailed molecular pathogen diagnostics and healthcare data can be captured and analyzed in near-real time, providing not only insights into disease outcomes and dynamics but also data-driven decision support for pandemic mitigation resource allocation and policymaking. In this regard, the COVID-19 pandemic stands in contrast to the 1918 influenza pandemic, in which World War I-associated disruptions and the nascent state of the public health surveillance infrastructure impeded consistent collection of morbidity and mortality statistics across the United States. Notwithstanding such limitations, available data from the 1918 influenza pandemic point toward health disparities in influenza outcomes in Black Americans versus White Americans, presaging those being observed in the present-day COVID-19 pandemic and suggesting that health inequity across racial (and socioeconomic) lines has remained a continuing concern in the United States well into the 21st century.

During the 1918 influenza pandemic, compared to the White population in the United States, Black Americans exhibited lower influenza incidence and morbidity rates but higher mortality rates [1, 2]. Various hypotheses have been advanced to explain this observation. Some have argued that race itself could have explained these differences, whereby supposed race-associated biological differences may have somehow contributed to heightened immunity of Black individuals to influenza infection [3]. Others have hypothesized that structural racism, rather than race per se, was largely responsible for racial health disparities observed during the 1918 influenza pandemic. For example, if the living and social conditions experienced by Black Americans of that era had predisposed them to acquiring infection with earlier, less virulent strains of influenza, then any partial immunity that may have been acquired might have lowered attack rates of the subsequent 1918 influenza strain amongst Black persons [4]. For those Black individuals who did become infected with the 1918 influenza strain, it might also be argued that their observed higher mortality rates could have been in part due to Black Americans’ generally poorer health status and reduced access to equitable health care, both of which stemmed from the consequences of structural racism.

In the intervening century since the 1918 influenza pandemic, despite an overall demographic shift toward a more pluralistic and multi-racial society in many regions of the United States, racial and ethnic disparities continue to persist in various domains, including in health, and are evident not just for Black Americans but for Americans belonging to a range of racial and ethnic minorities. During the current coronavirus pandemic, Black and Hispanic individuals have tested positive for COVID-19 at higher rates than White individuals [5, 6, 7, 8], and COVID-19-related hospitalizations and deaths have disproportionately affected American Indian, Asian, Black, and Hispanic persons [9, 10, 11, 12]. Despite widespread recognition of racial health disparities dating back more than a century, considerable disagreement and uncertainty remain as to whether the principal cause of health inequities is more firmly rooted in race or in racism.

An alternative framework for understanding racial/ethnic health disparities might consider contributory factors as occupying one of two domains, each broadly defined but inherently intersectional. The first domain would comprise *individual-specific* factors that correlate with a person’s susceptibility to and manifestation of disease, which may reflect allelic, epigenetic, and behavioral variations that are influenced by an individual’s race/ethnicity, age, gender, health status, genetic background, ancestry, culture, beliefs, and health-related practices. The second domain would encompass *contextual, group-specific* factors that reflect multiplex systemic, historical, legal, economic, social, interpersonal, and structural racism-related impacts on minority and vulnerable populations, including with respect to their attributes of health, access to health care, and experience of health care delivery.

Arguably, neither nature nor nurture alone can explain all observed differences in health measures based on race and ethnicity. On the one hand, the phenotypic expression of disease is dependent not just on intrinsic genomic determinants but also by multiple epigenetic and environmental externalities, including age, gender, co-morbidities, diet, substance use, exposures, and health-related behaviors, all of which are intimately tied to an individual’s life experiences and social context. Conversely, an individual’s environment and experience of racism may have differential effects on health depending on that person’s age, gender, medical status, genetics, and other biomedical predispositions as well as individual-specific health-related behaviors and beliefs.

Accordingly, the debate then properly shifts from argumentation as to whether race alone or racism alone can explain all observed health equity differences to a more nuanced discussion of the relative contribution and impact of a spectrum of factors across biomedical, behavioral, social, and demographic domains on varying aspects of individual and group health and associated measures of health equity. This subtle but critical distinction is key not only to creating a more inclusive framework for understanding the underlying causes of health disparities but also essential for appropriately crafting policy, wisely allocating resources, and effectively implementing measures to holistically address racial and ethnic health inequities.

To the extent that the consequences of slavery, Jim Crow laws, oppressive policing policies, inequitable housing conditions, and discriminatory immigration and refugee policies continue to perpetuate mutually reinforcing, systemically rooted societal inequalities in education, employment, advancement opportunities, and income/wealth; and to the degree to which the resulting reinforcement of stereotyped treatment and resource maldistribution translate into poorer mental and physical wellness and reduced access to health care, then systemic and structural racist practices can be reasonably viewed as important contributors to racial and ethnic health disparities. Efforts to reduce health disparities must then include addressing both structural and interpersonal racism, as well as their immediate and secondary consequences for oppressed, marginalized, and vulnerable groups.

By extension, to the degree that an individual’s biomedical makeup and/or behavior can directly or indirectly influence the incidence, transmission, manifestation, and treatment of disease; and to the degree to which such factors are intimately related to race/ethnicity, whether through ancestry-related genetic/allelic variations or health-related behaviors associated with a person’s race/ethnicity (e.g., diet, habits, household composition, health beliefs, health practices, etc.), then measures targeted at improving health, promoting health-conscious behaviors, and providing culturally sensitive and contextually appropriate care become instrumental to devising comprehensive and effective approaches to closing health equity gaps.

In order to better understand the relative contributions of biomedical, racial/ethnic, and socioeconomic factors to health disparities in the COVID-19 pandemic, we examined COVID-19 test positivity and hospitalization rates in a racially, ethnically, and socioeconomically diverse region of the United States, the San Francisco Bay Area. The Bay Area is of particular value for conducting such a study due to its pluralistic racial and ethnic composition and the region’s relative decoupling of race/ethnicity and socioeconomic status as compared to many other US metropolitan areas, particularly for Asian individuals [9].

We previously reported an initial analysis of racial and ethnic differences in COVID-19 test positivity and hospitalization rates among patients of a large, multi-hospital, multi-clinic academic health care system in the San Francisco Bay Area [9]. We found that higher social vulnerability, Black race, and Hispanic ethnicity were associated with a higher rate of COVID-19 positivity, whereas COVID-19 hospitalization and morbidity rates were highest among Asian patients. As members of the largest racial group, White persons tallied large numbers of COVID-19-related deaths, but Hispanic individuals who died of COVID-19 had the lowest mean age and thus suffered the highest average number of years of life lost.

In the present study, we conducted a more detailed analysis of the contributing factors to COVID-19 positivity and hospitalization among the same study population, specifically extending our investigation to include a number of individual-specific biomedical correlates of health. We then developed logistic regression models to examine the relative contribution of racial/ethnic, biomedical, and socioeconomic factors to COVID-19 outcomes.

## Methods

In order to mitigate the potentially confounding association between race/ethnicity and socioeconomic status in many areas of the United States, we undertook our study in the San Francisco Bay Area, a largely urban/suburban region characterized by both high racial/ethnic diversity and high socioeconomic diversity. The Bay Area population of 7.8 million residents (2020 US Census) is majority non-White, with a racial/ethnic demographic composition consisting of 36% Whites, 6% Blacks, 28% Asians, 24% Hispanics, and 6% Other (2020 US Census). In addition, this metropolitan region also demonstrates marked income inequality, as evidenced by an 11-fold income difference between households in the 90th and the 10th income percentiles and the observation that fully one-third of the households are characterized as very low income [13, 14]. The Bay Area Asian population is particularly diverse, both ethnolinguistically as well as socioeconomically. According to the 2000 Census, 112 languages are spoken in the Bay Area, with more than half of the top 10 languages spoken in San Francisco being Asian languages [15]. Bay Area Asian individuals are a particularly socioeconomically diverse group. As a population, Asian persons are evenly distributed across the income spectrum, with 31% of the group occupying the very low-income category and 36% the high-income category [16, 14].

### Data

Under an institutional review board (IRB)-approved protocol (IRB #20-30545), we analyzed the University of California, San Francisco (UCSF) COVID-19 Electronic Health Record (EHR) Data for Research. UCSF Health is a large, multi-hospital, multi-clinic academic medical health system with locations in five of the nine counties of the greater San Francisco Bay Area. The database included the complete records of all UCSF Health patients who had undergone reverse transcriptase polymerase chain reaction (RT-PCR) testing, either as an outpatient or an inpatient, from January 1, 2020 to December 31, 2020, inclusive. In addition, under an IRB-approved protocol, we geocoded each patient’s address and attached US Census Bureau data on demographics and socioeconomic characteristics corresponding to the smallest available census unit (census block or census tract) of the patient’s residence. All other personally identifiable data were redacted to preserve confidentiality of protected health information.

The complete data set was censored to include only records of patients who met all three of the following criteria: residency in one the nine greater San Francisco Bay Area counties (Alameda, Contra Costa, Marin, Napa, San Francisco, San Mateo, Santa Clara, Solano, and Sonoma); self-reporting of a single race or ethnicity; and completion of at least one SARS-CoV-2 RT-PCR test in calendar year 2020, during the pre-vaccination phase of the pandemic. This time period was chosen to exclude the influence on clinical outcomes of vaccinations, none of which could have been completed until January 2021 for the earliest vaccinated individuals, a subset that constituted < 1% of the total study population. Since RT-PCR testing was not widely available during the first several months of 2020, relatively few tests were recorded in January 2020 and February 2020, but the number of tests increased steadily thereafter, starting in March 2020. The earliest test in the data set was performed on January 4, 2020, and the latest test result was performed December 31, 2020.

Race/ethnicity was self-reported and classified as non-Hispanic Black (hereafter, Black), non-Hispanic White (hereafter, White), non-Hispanic Asian (hereafter Asian), or Hispanic or Latino (hereafter Hispanic). Because the small number of observations would preclude adequate statistical analysis of the Native Hawaiian or Other Pacific Islander group (< 1%), these patients were aggregated with the Asian group. Similarly, we did not conduct a separate analysis for American Indian or Alaska Native patients (< 1%) or for patients self-reporting as multi-racial (2.6%) because their small numbers in our data set preclude a statistically meaningful analysis for these groups. A number of individuals did not self-report a race or ethnicity (8.4%). These patients, like the American Indian or Alaska Native and multi-racial patients were included in the analyses of all patients but were excluded from the race-specific analyses.

Outcome measures included COVID-19 test (SARS-CoV-2 RT-PCR) positivity and COVID-19-related hospitalization. Hospitalization for COVID-19 was defined by a non-procedural hospitalization for more than 24 hours with an admitting diagnosis of COVID-19. For hospitalized patients, each record was examined for the presence of a set of biomedical factors/co-morbidities that have been reported in one or more previous studies to be associated with COVID-19 morbidity or mortality [17, 18, 19, 20, 21, 22]. Conditions were defined by either the recording of a corroborative ICD-9 or ICD-10 diagnostic code in the EHR and/or by a laboratory value during the hospitalization that was consistent with the diagnosis. The conditions identified and the manner in which they were identified are as follows:

- Diabetes mellitus was defined by ICD-10 diagnosis of Type I diabetes mellitus (E10) or Type II diabetes mellitus (E11); by ICD-9 codes 249 or 250; and/or by any of the following abnormal laboratory values: blood hemoglobin A1C *>* 6.4%, serum glucose (either random or after an oral glucose tolerance test) *>* 200 mg/dL, or fasting serum glucose *>* 125 mg/dL. A subgroup of those with poorer glycemic control was defined by having a hemoglobin A1C *>* 7.5%;
- Kidney disease was defined by an ICD-10 code indicating chronic kidney disease (N18) and/or dialysis status (Z99.2); by ICD-9 codes 585.1, 585.2, 585.3, 585.4, 585.5, 585.6, and/or 585.9; and/or by a serum creatinine level that was above the normal range for the patient’s age and gender (e.g., for adults < 60 years, serum creatinine *≥* 1.3 mg/dL in male patients and *≥* 1.1 mg/dL in female patients);
- Hypertension was denoted by ICD-10 diagnoses I10, R03.0, I16.0, I11, I15, I12, I50.2, and/or I50.4; ICD-9 code 401; and/or by at least two diastolic blood pressure measurements recorded on two separate days during hospitalization that exceeded 100 mm Hg;
- Dyslipidemia was defined by ICD-10 diagnostic code E78.5, ICD-9 code 272, and/or serum cholesterol *≥* 240 mg/dL;
- Obesity was defined by ICD-10 code E66, ICD-9 code 278, and/or calculated Body Mass Index (BMI) *>* 30 kg/m^2^. Severe obesity was defined by ICD-10 codes E66.01, E66.02, and/or Z68.4; ICD-9 code 278.01, and/or BMI *>* 35 kg/m^2^;
- Chronic Obstructive Pulmonary Disease (COPD) was defined by ICD-10 codes J42, J43, and/or J44; or ICD-9 codes 491.21, 491.22, 493.21, 493.22, 491.9, 492.8, and/or 492.0;
- Asthma was defined by ICD-10 code J45 or ICD-9 code 493;
- Cardiac disease was identified by ICD-10 codes I20-I25, I40, I48, I49, and/or I50; or ICD-9 codes 410, 411, 413, 414, 422, 428, 427, 429, and/or 440;
- Liver disease was defined by any of the ICD-10 codes between K70–K77, inclusive; or by ICD-9 codes 571, 572, and/or 573;
- Coagulation disorder was recorded for any ICD-10 code between D66–D69, inclusive, or for ICD-9 code 286;
- Cerebrovascular disorder was identified by ICD-10 codes G45, G46, H34.0, I6x, I97.81, and/or I97.82; or by ICD-9 codes 437 and/or 438;
- Cancer was defined by any of the ICD-10 codes between C01–Cxx, inclusive, in which “xx” denotes any two digit number; or by ICD-9 codes 140-199, inclusive; and
- Smoking status, past or present, was designated by ICD-10 diagnoses F17, U07.0, T65.2, V15.82, Z71.6, Z72.0, or Z87.891; ICD-9 code 305.1 or V15.82; and/or by current smoking status or a past smoking history as recorded in the Social History section of the EHR.

Note that in order to remove any confounding effects of conditions that may have developed due to complications of COVID-19, only those conditions that were diagnosed prior to hospitalization were included. The exceptions were diabetes and hypertension, both of which are associated with relatively high rates of under-diagnosis. So as not to exclude newly diagnosed cases of either condition, we allowed these diagnoses to be established within three days of hospitalization.

We examined both individual-specific demographic factors, including age, gender, and recorded medical insurance carrier, as well as context-specific factors, including a number of metrics associated with the patient’s place of residence such as average household size, geocoded to the census block level, and the Centers for Disease Control Social Vulnerability Index (CDC SVI), geocoded at the census tract level. The CDC SVI is a validated index that incorporates four different sub-measures: socioeconomic factors, household composition and disability status, minority status and primary language spoken, and housing type and primary transportation modality employed [23]. Health insurance status was denoted as “Medicaid” if at least one of the payors was Medicaid, MediCal (the California state Medicaid program), or the equivalent, or if the listed payor was listed as “charity care,” “no insurance carrier,” or the equivalent. Health insurance status was labeled as “private insurance” if at least one listed payor was a commercial insurance carrier.

Two sets of logistic regression analyses were performed. In the first analysis, all patients who underwent COVID-19 testing were included, and the dependent variable was COVID-19 test positivity. In the second, the data were restricted to those patients who tested positive for COVID-19, and the dependent variable indicated whether the patient was hospitalized for COVID-19. For the independent variables in the test positivity analysis, we present results for our model in which only sociodemographic variables were included. We then performed a subsequent analysis in which both individual-specific biomedical/health factors and contextual sociodemographic variables were included. For the hospitalization analysis, both health conditions and sociodemographic aspects were included as independent variables. Statistical significance was assessed at the 0.05-level. If an independent variable was found to be statistically significant, the adjusted odds ratio for that variable was computed, along with its associated 95% confidence interval.

## Results

### COVID-19 Positivity

Differences in the raw COVID-19 positivity rates were observed between the different racial/ethnic groups. In particular, rates of COVID-19 positivity were highest for Hispanic patients (11.8%), followed in descending order by Black patients (5.4%), Asian patients (3.7%), and White patients (2.5%). The difference between Hispanic and White patients was more than 4.5-fold. A logistic regression analysis was conducted to assess the contribution of various factors to these observed differences in rates of test positivity.

Five separate models were analyzed: one including all patients, and four others restricted to patients from one of the four racial/ethnic groups (Black, White, Asian, and Hispanic). Table 2 in the Appendix displays the full regression results. Figure 1 shows the adjusted odds ratios (ORs) along with their associated 95% confidence intervals for only the variables that were statistically significant at the 0.05-level.

**Figure 1:**
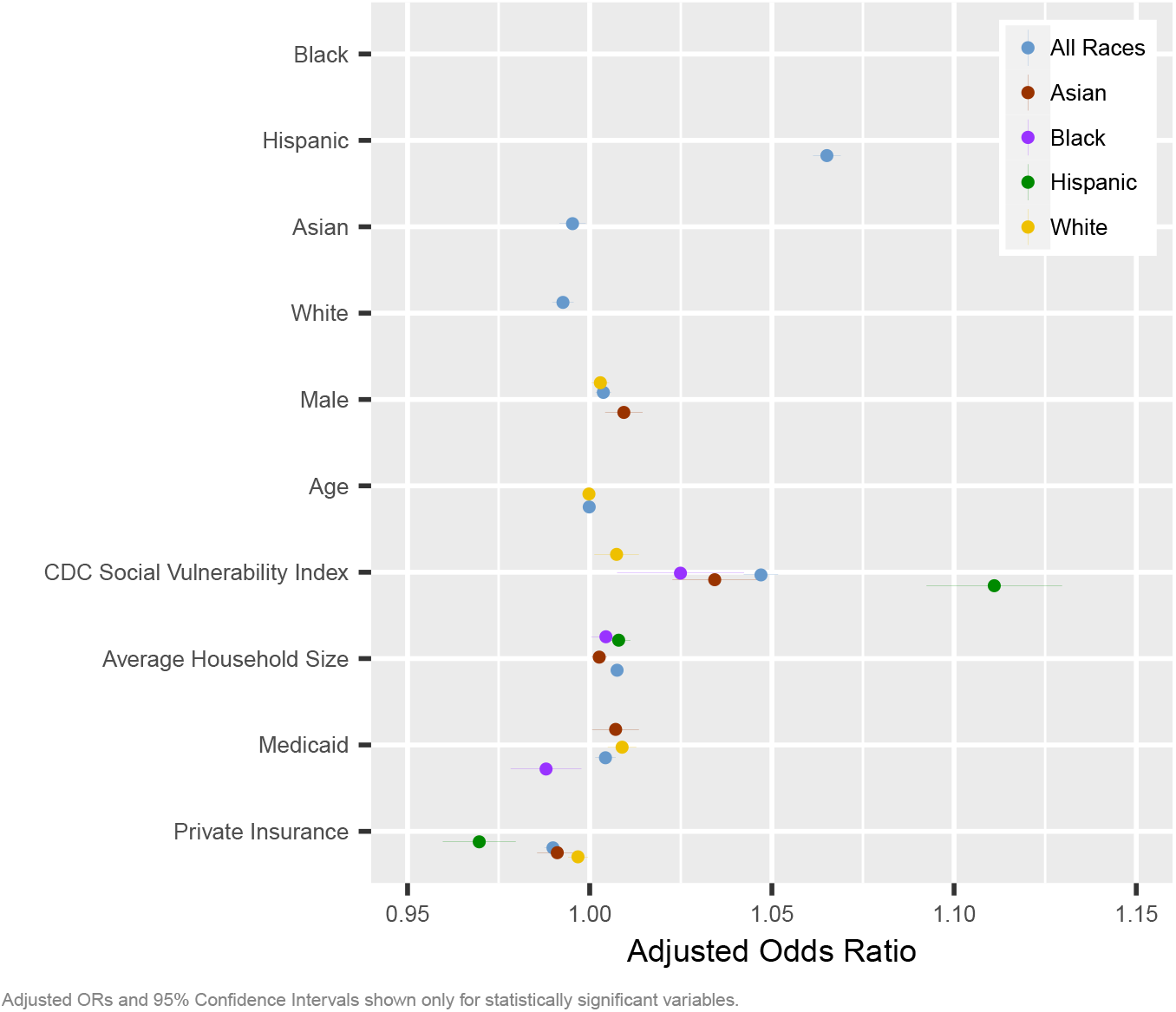
COVID-19 Positivity Risk Factors.

The higher test positivity rate for the Hispanic group and the lower test positivity rates for Asian and White populations remained statistically significant even after controlling for age, gender, medical insurance coverage, average household size, and CDC Social Vulnerability Index. Thus, in our study population, for all groups except the Black population, the association of race/ethnicity and COVID-19 test positivity was statistically significant even after controlling for other demographic or socioeconomic variables.

Medicaid patients tended to test positive at higher rates, whereas those with private insurance tended to test positive at lower rates. A small positive relationship was noted between living in an area with larger median household sizes and the likelihood of testing positive for COVID-19. These effects, while statistically significant, were relatively small in magnitude. A substantially larger effect on testing positivity was seen for those who resided in areas with higher social vulnerability, particularly among Hispanic individuals. The odds of a positive test were 11% higher among Hispanic individuals who resided in the most socially vulnerable census tracts versus Hispanic persons who resided in the least vulnerable census tracts. While this relationship between higher social vulnerability and higher likelihood of testing positive was observed for all four racial/ethnic groups, the risk was highest for those whose ethnicity was Hispanic.

It is possible that people who resided in poorer living conditions may have been more susceptible to testing positive for COVID-19 because they were in generally poorer health. To explore this hypothesis, we performed a logistic regression model that included not just sociodemographic variables, but also biomedical risk factors (including diabetes, obesity, dyslipidemia, hypertension, cardiac disease, cerebrovascular disease, kidney disease, chronic obstructive pulmonary disease, asthma, smoking history, coagulopathy, and cancer). Interestingly, in this secondary analysis, while some health conditions reached the threshold of being statistically significant, the odds ratios were almost all less than 1.01 (i.e., 1% increased risk) and furthermore, most of the effects were slightly negative (i.e., protective) rather than positive. Accordingly, on the whole, the examined biomedical factors had no clinically meaningful impact on the likelihood of testing positive for COVID-19. Aside from study-specific power limitations to detect such an effect, it is possible that any such effect, even if present, could have been offset by other factors, such as more medically vulnerable individuals practicing more stringent behavioral mitigation measures such as mask wearing and avoiding social gatherings, in order to avoid contracting COVID-19. Regardless, whether biomedical conditions are considered or not, the results of both sets of regression models indicate that race/ethnicity and increased social vulnerability were statistically significant indicators of COVID-19 test positivity.

### COVID-19 Hospitalization

Of the 7,280 individuals testing positive for COVID-19, 696 (6.5%) were hospitalized at UCSF Health. As noted previously [9], although Asian patients had the second-lowest rates of testing positive for COVID-19 amongst the four racial/ethnic groups, Asian patients demonstrated the highest rate of hospitalization. To investigate this observation further, a logistic regression was performed that included race/ethnicity, sociodemographic factors, and biomedical risk factors as independent variables, and COVID-19 hospitalization as the dependent variable. Inclusion criteria included all patients who tested positive for COVID-19. Five models were run: one including all individuals and one model each for the four studied racial/ethnic groups. Full regression results are presented in the Appendix (Table 3). Figure 2 shows the summary results in graphical form, displaying, in descending rank order, only those variables found to have a statistically significant effect on the risk of hospitalization.

**Figure 2:**
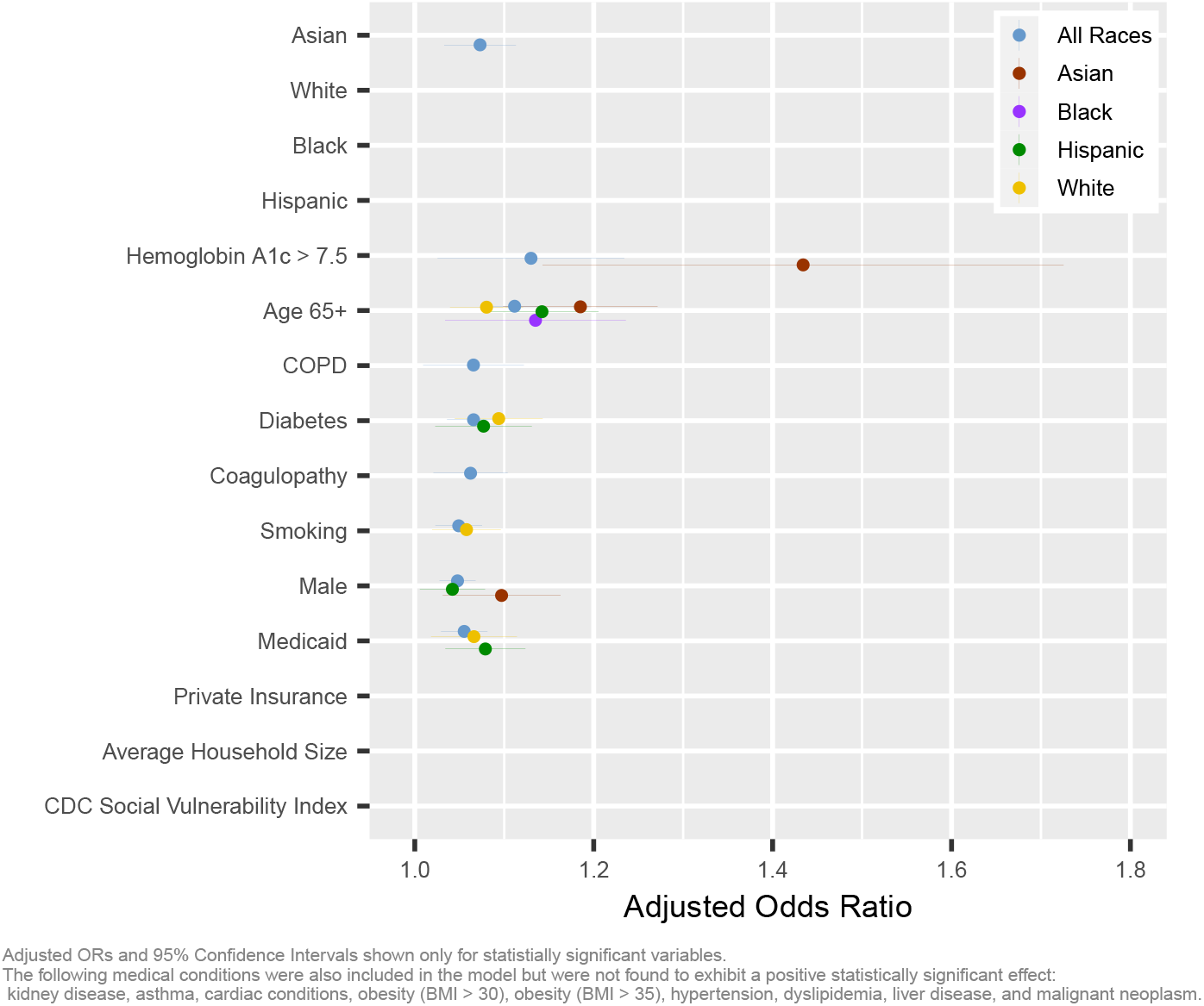
COVID-19 Hospitalization Risk Factors.

Asian race conferred an increased and independent risk of COVID-19 hospitalization, even after controlling for biomedical and other sociodemographic factors. None of the other racial/ethnic groups was associated with either an increased or decreased risk of hospitalization.

Whereas the CDC Social Vulnerability Index (CDC SVI) was observed to be a statistically significant predictor of COVID-19 test positivity, neither the CDC SVI nor average household size had a statistically significant relationship with COVID-19 hospitalization. Only insurance status was found to have a statistically significant relationship, with Medicaid or no health insurance elevating the risk of hospitalization. Again, in contradistinction to the finding that biomedical factors had little to no effect on COVID-19 test positivity, a number of health conditions were found to be independent and statistically significant predictors of COVID-19 hospitalization (Figure 2). For the entire population, conditions that were associated with COVID-19 hospitalization included the following, in descending order of importance: higher severity diabetes with hemoglobin A1C *>* 7.5%, age ≥ 65 years, chronic obstructive pulmonary disease and diabetes (without respect to severity), coagulopathy, smoking, and male gender.

On an unadjusted basis, the presence of diabetes conferred a ∼26% elevation in risk of COVID-19 positivity amongst Asian patients, whereas that risk was elevated by only ∼12% when diabetes was present in White patients (Table 1). There was no appreciable increase in unadjusted COVID-19 positivity rates when diabetes was present in either Black or Hispanic patients.

**Table 1:**
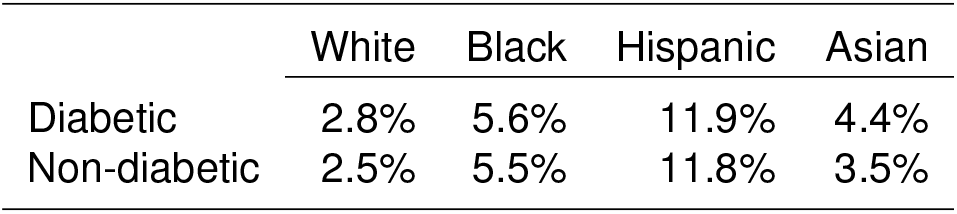
Rates of COVID-positivity for Diabetic vs Non-Diabetic Patients, According to Race/Ethnicity.

Age and diabetes increased hospitalization risk to varying degrees, depending on a patient’s race/ethnicity. Asian patients 65 years and older were at somewhat higher risk for hospitalization than individuals age 65 years and older from other racial/ethnic groups. Higher glycohemoglobin levels (*>* 7.5%), indicative of poorer glycemic control, were associated with a moderately increased hospitalization risk (adjusted odds ratio of 1.4), especially for Asian diabetics. For White diabetics, the adjusted odds ratio (1.3) for higher glycohemoglobin levels (*>* 7.5%) was *p*-value was 0.057 (Table 3), a level close to the predefined threshold of statistical significance. Less severe diabetes was also a risk factor for hospitalization, though it conferred a more mild risk. The 7.7% increased risk of hospitalization for Hispanic diabetics and 9.4% increased risk for White diabetics were both significant at the 0.05-level, while an increased risk of 8.8% for Asian diabetics had a *p*-value of 0.051. Diabetes was not associated with an increase in hospitalization risk for Black patients.

The proportion of hospitalized COVID-19 patients with diabetes that had no diagnosis of diabetes prior to hospitalization was 49% among Asian patients, compared to 46% among white patients, 39% among Black patients, and 35% in Hispanic patients. Moreover, upon first diagnosis in the hospital, Asian patients had the highest mean hemoglobin A1C (7.5%), compared to 7.3% in Hispanic diabetics and 6.5% in both White and Black diabetics, indicating that previously undiagnosed Asian diabetics had the poorest glycemic control in comparison to newly diagnosed Hispanic, Black, and White diabetics.

## Discussion

### Divergence Between Correlates of COVID-19 Test Positivity and Hospitalization

In the cohort studied, race/ethnicity (except in Black individuals) and poorer socioeconomic status (e.g. having Medicaid insurance or no health insurance, residing in a household with a greater number of individuals, and/or having an increased social vulnerability score) were generally associated with an increased risk of testing positive for COVID-19, whereas an individual’s biomedical profile played little to no role in increasing the likelihood of testing positive. The CDC Social Vulnerability Index is comprised of a number of different components that could have contributed to an increased risk for COVID-19 exposure and/or susceptibility to infection, including lack of English proficiency, transportation challenges, residence proximate to environmental pollutants, and increased household density. Further investigation to separate these component of the CDC SVI would be needed to ascertain the relative impact of the SVI sub-components that contribute to COVID-19 transmission and/or susceptibility risk. Such a study would be helpful to design intervention and coordinated policy targeted at improving population health and addressing social determinants of health inequities, particularly for the most vulnerable subpopulations.

By contrast, for COVID-19 hospitalization, nearly the exact converse was observed. Almost all the statistically significant risk factors were in the biomedical domain. The highest risk factors were higher severity diabetes and advanced age (65 years or older), but other risk factors included coagulopathy, diabetes (regardless of severity), severe obesity, male gender, smoking, and asthma. Asian race, irrespective of biomedical and other sociodemographic factors, was an independent risk factor for hospitalization; Asian patients who tested positive for COVID-19 had more than twice the hospitalization rate of White patients who were COVID-19 positive. Of the other examined sociodemographic factors, only poorer health insurance status (Medicaid or no health insurance) was associated with a small increased risk of hospitalization. The other sociodemographic factors, including those included in the CDC Social Vulnerability Index, were not statistically significant predictors of hospitalization risk.

Although Black individuals tested positive at rates that were more than double that of White individuals and were hospitalized at rates second only to those of the Asian group, no additional effect of Black race was noted once sociodemographic and biomedical variables were taken into account. In other words, the excess risk attributable to being Black could be explained by other sociodemographic factors and biomedical predispositions. If these findings are corroborated, then efforts to alleviate COVID-19 health disparities for the Black population might best be directed towards addressing contributors to socially vulnerability and improving the health status of Black persons.

Hispanic ethnicity was an independent risk factor for COVID-19 positivity, irrespective of other sociodemographic factors, but was not significant for COVID-19 hospitalization. Furthermore, while increased social vulnerability was associated with a higher risk of COVID-19 positivity for all racial/ethnic groups, the magnitude of this effect was the highest for Hispanic patients. Although the reason(s) for this difference are unclear and warrant additional study, one potentially promising line of inquiry would be to consider whether this elevated risk is related to the significantly younger age profile of the Hispanic group. A report by the Pew Research Center found that in 2014, close to 60% of all Hispanic individuals were Millennials (ages 18 to 33) or younger. By comparison, half of the Black population, 46% of the U.S. Asian population, and 39% of White persons were of Millennial age or younger [24]. It is possible that younger Hispanic individuals are disproportionately represented in frontline (“essential worker”) jobs, in which the risk of COVID-19 transmission would be expected to be higher than for occupations allowing remote work from home. If these findings are validated, a focus on prevention in at-risk Hispanic subpopulations, including vaccination and behavioral, non-pharmaceutical interventions such as masking and social distancing could be helpful. It is also worth considering how the tasks and work environment of these essential worker jobs might be adjusted to mitigate coronavirus transmission and how health education, sick leave policy, and availability of affordable health care for essential workers might promote behaviors to reduce the risk of disease transmission in the workplace.

Asian individuals were distinct among the racial/ethnic groups in exhibiting a higher risk for COVID-19 hospitalization, even when adjusting for age, medical co-morbidities, and other sociodemographic characteristics. Again, further inquiry appears warranted since these data measures do not fully capture many of the potential barriers to health care often confronted by Asian patients, particularly those who are immigrants. Health care access and health care utilization in Asian individuals may be hindered, for instance, by language barriers, limited digital access, lack of ethnolinguistically concordant health care providers, preferences for non-Western traditional treatment approaches, and/or cultural differences in health care beliefs, behaviors, and practices. At least one study has observed that Asian immigrants utilize women’s health care services at lower rates than non-Hispanic White persons [25]. Indeed, immigrants form a markedly higher percentage of the US adult Asian population (71%) compared to the general US adult population (17%) [26]. Moreover, because Asian immigration to the US has historically been influenced by labor demands, many Asian immigrants, including older Asian adults, are in essential worker occupations that may pose a higher risk of COVID-19 exposure. One in five Asian adults aged 55 years and older work in frontline service jobs, compared to only 15% of the total US population [27]. Indeed, older Asian adults have a higher rate of hospitalization for COVID-19, even after controlling for other biomedical and sociodemographic factors. Further investigation is needed to examine whether the observed excess hospitalization rate in older Asian patients might be attributed in part to higher occupational COVID-19 exposure.

### Effect of Diabetes

The impact of diabetes in increasing the risk of COVID-19 positivity and COVID-19 hospitalization in Asian patients is noteworthy. Furthermore, the proportion of diabetics with previously undiagnosed diabetes was higher for Asian patients than for any other race/ethnicity, and the average hemoglobin A1C levels during hospitalization were again highest for Asian diabetics. A strong link between diabetes mellitus and COVID-19 hospitalization was reported in a 2020 Israeli study in which the only risk factor associated with an increased risk of COVID-19 hospitalization was severe diabetes with A1C *>* 9% [28]. In Asians, diabetes occurs more frequently in non-obese individuals [29] and at lower BMI [30]. A proposed explanation for these differences includes higher visceral adiposity for a given BMI in Asian individuals versus non-Asian individuals. Similar to what we observed, Menke et al. found that more than half of

Asian persons with diabetes were undiagnosed [31]. Other investigators have reported that diabetes is more frequently underdiagnosed in Asian and Hispanic individuals compared to White and Black persons [32]. Further study is needed to understand the mechanism by which diabetes contributes to excess morbidity risk in COVID-19. Efforts to mitigate excess risk of COVID-19 morbidity amongst Asians might focus on diagnosis and treatment of diabetes, but must also consider other factors that are specific to the Asian population (e.g., language and/or cultural issues that may affect willingness to seek health care and/or ability to access health care services).

### Study Limitations

Our study was conducted on the patient population of a large academic health care system in the San Francisco Bay Area during the pre-vaccination phase of the COVID-19 pandemic; accordingly, these specific findings may not be generalizable to populations in other settings or at different time points. Some differences in the racial and ethnic composition of the study cohort and that of the Bay Area were noted, possibly reflecting differences in the demographics of the counties closest to UCSF Health locations, variations in patient access to and willingness to seek care at UCSF Health care facilities, health insurance coverage–dictated limitations, and referral effects. Patients who lived a distance from a UCSF Health facility, and in particular individuals with higher social vulnerability and lower socioeconomic status, may have been less likely to seek care at UCSF Health and thus would been less likely to be included in the study population. Mitigating any such effects is the fact that, among large, regional health care systems in the San Francisco Bay Area during the study period, UCSF Health cared for a disproportionately greater share of uninsured and underinsured patients.

Errors in self-reported residence address data were corrected manually when possible (e.g., misspelled street names and transposed zip code digits) but records with missing street addresses or other unresolvable errors were excluded. These limitations may have introduced some systematic errors, such as underreporting of homeless individuals, but the magnitude of such errors is unknown.

The assignment of a socioeconomic vulnerability to all individuals within a given census tract, while a standard process in demographic studies, was a necessary limitation since the electronic health record did not include measures of sociodemographic data that are gathered by the US Census. Studies conducted in similar fashion to ours that examine just EHR data [33], by necessity, all rely on group-level rather than individual-specific measurements.

It was not possible to capture data from any individuals who may have moved out of the area or migrated to another health care system between the time of COVID-19 testing and hospitalization, so it is possible that the number of hospitalizations may have been underreported. However, there have been no published studies that would suggest that during the COVID-19 pandemic, any such effect would have been more likely to affect one racial and ethnic group over another or to preferentially affect individuals of higher or lower social vulnerability. Thus, even if a mild degree of undercounting had been present, the effects are unlikely to have been large or to have substantially altered the study findings.

## Conclusion

We observed that biomedical, racial/ethnic, and sociodemographic factors all contributed in varying but distinct ways to COVID-19 test positivity and hospitalization rates in a multiracial, socioeconomically diverse US metropolitan area. Hispanic ethnicity and increased social vulnerability correlated strongly with a higher likelihood of COVID-19 test positivity. Asian race, age, higher severity diabetes, older age, male gender, coagulopathy, smoking, and Medicaid/uninsured status all correlated with a higher likelihood of COVID-19-related hospitalization. Of the various biomedical co-morbidities, diabetes emerged as an important contributor to COVID-19 morbidity, particularly for Asian patients, who had the highest frequency of previously undiagnosed diabetes and the worst glycemic status.

Sociodemographic and group contextual factors generally appeared most important for influencing the risk of acquiring COVID-19 infection, whereas biomedical and individual-specific factors contributed to a higher likelihood of developing COVID-19-associated morbidity (i.e., hospitalization). Even so, there remained race/ethnicity-associated elevated risks (e.g., elevated test positivity rates for Hispanic individuals and increased hospitalization rates for Asian persons) that were not explained by the variables examined in this study. We have proffered a number of potential explanations for these race/ethnicity effects but further inquiry would be needed to elucidate the underlying causes and potentially guide appropriate mitigations for these effects.

We offer a new framework for evaluating and addressing causes of racial and ethnic health disparities, one that eschews a race versus racism dichotomy in favor of acknowledging a spectrum of factors that can influence health outcomes and resultant disparities, broadly classified into intersectional domains of *individual-specific* and *group contextual* factors. The former would include not only biomedical and genetic predispositions, but also epigenetic, behavioral, and other exogenous factors that operate at the individual level. The latter would include socioeconomic and demographic factors, including those that primarily affect a particular racial or ethnic group, suggesting possible underlying systemic or structural contributors to health disparities. Improving COVID-19 outcomes in the population overall and addressing racial and ethnic health disparities in particular will likely require a comprehensive, data-driven approach that includes not only diagnostic, vaccination-based, pharmacotherapeutic, and non-pharmaceutical interventions, but also strategies to target both individual-specific and group contextual factors that contribute to increased transmission, susceptibility, and severity of SARS-CoV-2-related disease.

## Data Availability

All data are available through UCSF after IRB approval.

## Appendix

**Table 2:** Logistic Regression Coefficients for COVID-19 Positivity.

|  | All Races | Black | White | Asian | Hispanic |
| --- | --- | --- | --- | --- | --- |
| Intercept | 0.0202* (1.0205) | 0.0451* (1.0461) | 0.037* (1.038) | 0.019* (1.019) | 0.058* (1.060) |
| Age | -0.0002* (0.9998) | -0.0002 | 0.000* (1.000) | 0.000 | 0.000 |
| Male | 0.0037* (1.0037) | 0.0022 | 0.003* (1.003) | 0.009* (1.009) | 0.003 |
| Asian | -0.0048* (0.9952) |  |  |  |  |
| Black | -0.0007 |  |  |  |  |
| White | -0.0073* (0.9927) |  |  |  |  |
| Hispanic | 0.0631* (1.0652) |  |  |  |  |
| Medicaid | 0.0044* (1.0044) | -0.0121* (0.9880) | 0.009* (1.009) | 0.007* (1.007) | -0.007 |
| Private Insurance | -0.0102* (0.9898) | -0.0011 | -0.003* (0.997) | -0.009* (0.991) | -0.031* (0.970) |
| Average Household Size | 0.0073* (1.0074) | 0.0044* (1.0044) | -0.001 | 0.003* (1.003) | 0.008* (1.008) |
| CDC Social Vulnerability Index | 0.0459* (1.0470) | 0.0246* (1.0249) | 0.007* (1.007) | 0.034* (1.034) | 0.105* (1.111) |
| Total Testing | 133,390 | 10,550 | 72,774 | 23,880 | 22,744 |

Logistic regression coefficients shown in first column
Adjusted odds ratios for statistically significant variables shown in second column
\* $p < 0.05$

**Table 3:**
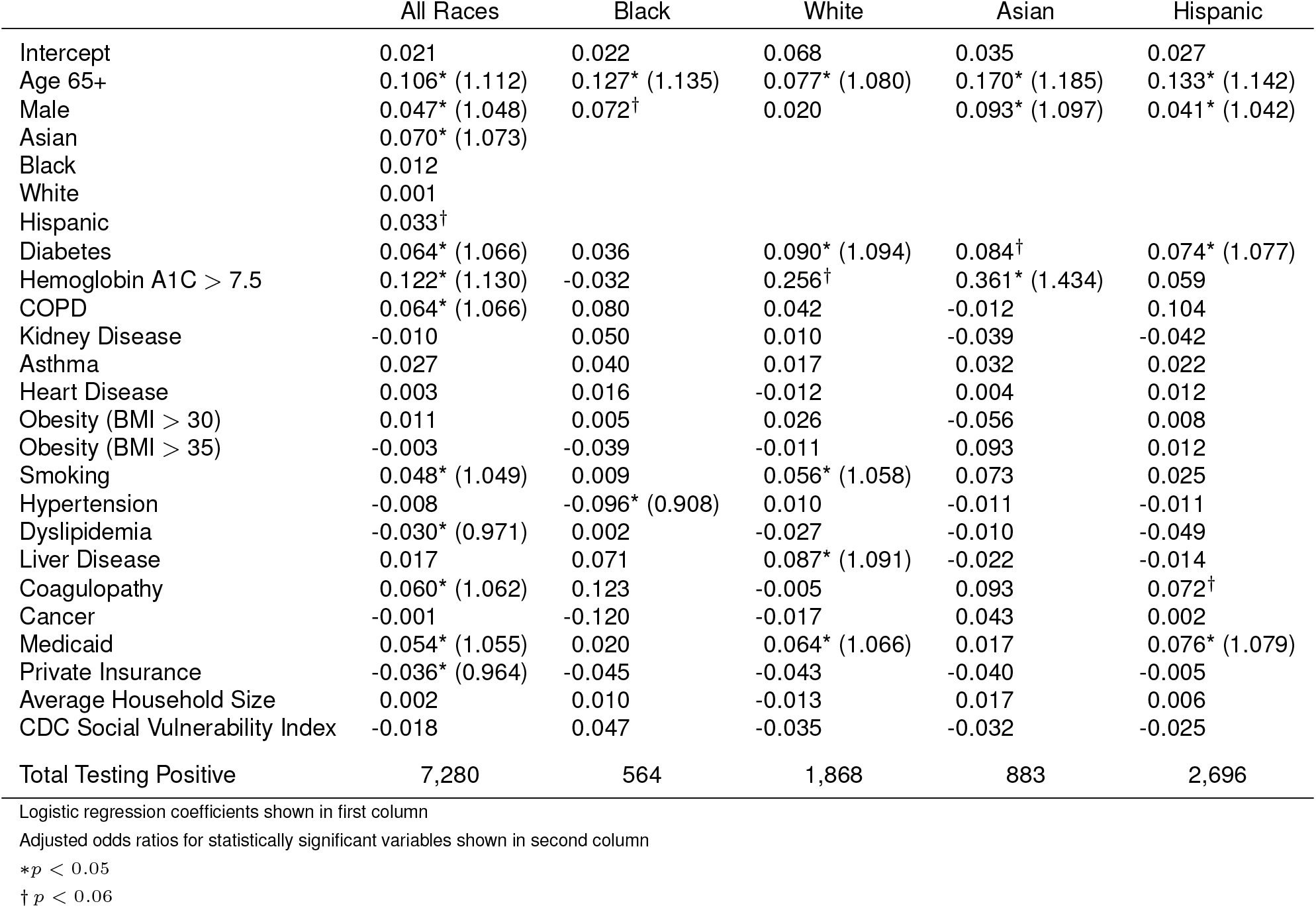
Logistic Regression Coefficients for COVID-19 Hospitalization.

|  | All Races | Black | White | Asian | Hispanic |
| --- | --- | --- | --- | --- | --- |
| Intercept | 0.021 | 0.022 | 0.068 | 0.035 | 0.027 |
| Age 65+ | 0.106* (1.112) | 0.127* (1.135) | 0.077* (1.080) | 0.170* (1.185) | 0.133* (1.142) |
| Male | 0.047* (1.048) | 0.072† | 0.020 | 0.093* (1.097) | 0.041* (1.042) |
| Asian | 0.070* (1.073) |  |  |  |  |
| Black | 0.012 |  |  |  |  |
| White | 0.001 |  |  |  |  |
| Hispanic | 0.033† |  |  |  |  |
| Diabetes | 0.064* (1.066) | 0.036 | 0.090* (1.094) | 0.084† | 0.074* (1.077) |
| Hemoglobin A1C > 7.5 | 0.122* (1.130) | -0.032 | 0.256† | 0.361* (1.434) | 0.059 |
| COPD | 0.064* (1.066) | 0.080 | 0.042 | -0.012 | 0.104 |
| Kidney Disease | -0.010 | 0.050 | 0.010 | -0.039 | -0.042 |
| Asthma | 0.027 | 0.040 | 0.017 | 0.032 | 0.022 |
| Heart Disease | 0.003 | 0.016 | -0.012 | 0.004 | 0.012 |
| Obesity (BMI > 30) | 0.011 | 0.005 | 0.026 | -0.056 | 0.008 |
| Obesity (BMI > 35) | -0.003 | -0.039 | -0.011 | 0.093 | 0.012 |
| Smoking | 0.048* (1.049) | 0.009 | 0.056* (1.058) | 0.073 | 0.025 |
| Hypertension | -0.008 | -0.096* (0.908) | 0.010 | -0.011 | -0.011 |
| Dyslipidemia | -0.030* (0.971) | 0.002 | -0.027 | -0.010 | -0.049 |
| Liver Disease | 0.017 | 0.071 | 0.087* (1.091) | -0.022 | -0.014 |
| Coagulopathy | 0.060* (1.062) | 0.123 | -0.005 | 0.093 | 0.072† |
| Cancer | -0.001 | -0.120 | -0.017 | 0.043 | 0.002 |
| Medicaid | 0.054* (1.055) | 0.020 | 0.064* (1.066) | 0.017 | 0.076* (1.079) |
| Private Insurance | -0.036* (0.964) | -0.045 | -0.043 | -0.040 | -0.005 |
| Average Household Size | 0.002 | 0.010 | -0.013 | 0.017 | 0.006 |
| CDC Social Vulnerability Index | -0.018 | 0.047 | -0.035 | -0.032 | -0.025 |
| Total Testing Positive | 7,280 | 564 | 1,868 | 883 | 2,696 |

Logistic regression coefficients shown in first column
Adjusted odds ratios for statistically significant variables shown in second column
\* $p < 0.05$
† $p < 0.06$

## Statements and Declarations

### Author Contribution

**Concept and design:** Both authors

**Acquisition, analysis, or interpretation of data:** Both authors

**Drafting of the manuscript:** Both authors

**Critical revision of the manuscript for intellectual content:** Both authors.

### Competing Interests

The authors have no relevant financial or non-financial interests to disclose.

### Funding

This research was supported in part by unrestricted grants to DGH from All May See (formerly That Man May See) and Research to Prevent Blindness.

### Compliance with Ethical Standards

This study was approved by the University of California San Francisco Human Research Protection Program Institutional Review Board (IRB #2030987. Reference #324788).

### Conflicts of interest

The authors declare that they do not have a conflict of interest.

## Acknowledgments

The authors wish to thank Ashly Dyke for assistance with the literature review. This work used the Extreme Science and Engineering Discovery Environment (XSEDE), which is supported by National Science Foundation grant number ACI-1548562. Computing resources were granted through the HPC Consortium for access to Pittsburgh Supercomputer Center’s Bridges, Bridges-2, and Bridges-AI.

## Notes

### Competing Interest Statement

The authors have declared no competing interest.

### Author Declarations

University of California San Francisco Human Research Protection Program Institutional Review Board IRB #20-30987 (Reference #314215)

